# Positivity in serum MMP-3 after clinical remission or low disease activity at 52 weeks leads to future joint destruction in patients with rheumatoid arthritis

**DOI:** 10.1101/2024.04.14.24305643

**Authors:** Hiroe Konishi, Mai Morimoto, Kosaku Murakami, Hideo Onizawa, Akira Onishi, Takayuki Fujii, Koichi Murata, Masao Tanaka, Akio Morinobu, Masayoshi Nakano, Masahiro Koshiba

**Affiliations:** Department of Clinical Laboratory Medicine, Hyogo Medical University School of Medicine, Hyogo, Japan; Division of Clinical Immunology and Cancer Immunotherapy, Center for Cancer Immunotherapy and Immunobiology, Graduate School of Medicine, Kyoto University, Kyoto, Japan; Department of Advanced Medicine for Rheumatic Diseases, Graduate School of Medicine, Kyoto University, Kyoto, Japan; Department of Orthopaedic Surgery, Graduate School of Medicine, Kyoto University, Kyoto, Japan; Department of Rheumatology and Clinical Immunology, Graduate School of Medicine, Kyoto University, Kyoto, Japan

**Keywords:** Rheumatoid arthritis, matrix metalloproteinase-3, C-reactive protein, modified van der Heijde total sharp score, joint destruction

## Abstract

**Introduction:** This study aimed to evaluate whether a long-term increase in serum matrix metalloproteinase-3 (MMP-3) levels leads to joint destruction in rheumatoid arthritis (RA) patients with negative serum C-reactive protein (CRP) values after methotrexate (MTX) therapy.

**Methods:** Patients with RA (n = 182) whose CRP values became negative due to MTX therapy were divided into two groups based on their MMP-3 positivity at the end of the observation period, and the 1-year progression of joint destruction was retrospectively compared. Radiological joint destruction was assessed using the modified van der Heijde total sharp score (mTSS).

**Results:** Among 109 (MMP-3(−), n = 63; MMP-3(+), n = 46) patients who achieved low disease activity or clinical remission (28 joint disease activity score erythrocyte sedimentation rate < 2.6), joint destruction (ΔmTSS ≥ 0.5) progressed in 24.6% and 48.9% of the MMP-3(−) and MMP-3(+) groups (p < 0.01), respectively. Prednisolone-induced increases in serum MMP-3 levels also resulted in joint destruction.

**Conclusion:** To prevent progressive joint destruction, the target MMP-3 value is 49.7 ng/mL in female patients, which is below the current MMP-3 cutoff value of 59.7 ng/mL. Residual MMP-3 activity may lead to the progression of joint destruction in patients with RA, even after CRP normalization by successful treatment with MTX.

## INTRODUCTION

Rheumatoid arthritis (RA) is a systemic autoimmune disease characterized by chronic synovitis and joint destruction, which leads to disability [1]. According to current recommendations, achieving clinical remission is the therapeutic target for patients with RA, with low disease activity (LDA) considered the best possible alternative. A treat-to-target strategy should be employed when treating patients with RA, and treatment decisions should be based on disease activity and other patient factors, such as comorbidities and the progression of structural damage [2, 3].

Methotrexate (MTX) is recommended as the first-line drug for the initial treatment of RA [4, 5]. MTX remains the anchor drug in RA. MTX is not only an efficacious conventional synthetic (cs) disease-modifying antirheumatic drug (DMARD) but is also the basis for combination therapies, either with prednisolone (PSL) or with other csDMARDs, biological DMARDs (bDMARDs), or targeted synthetic DMARDs (tsDMARDs) [6]. While serum CRP values often become negative after MTX therapy, joint destruction can still progress in some cases [7].

Matrix metalloproteinases (MMPs) are a family of enzymes that catalyze extracellular matrix degradation. Most MMPs are secreted as inactive preproteins that are activated when cleaved by extracellular proteinases [8]. MMP-3 is a proteinase secreted by synovial fibroblasts and chondrocytes in joints. In RA, joint destruction can be accelerated by active MMP-3, and serum MMP-3 levels in RA have been well evaluated as an indicator of disease activity. In RA, the level of MMP-3 within the joints is markedly higher than that of other MMPs [9]. This high MMP-3 level is believed to mediate joint destruction in patients with RA [10, 11].

Increases in serum MMP-3 levels are observed from early to advanced stages in 80%–90% of patients with RA. Serum MMP-3 levels in patients with RA reflect the degree of synovial proliferation and may serve as a prognostic indicator of disease progression, particularly in early RA onset [12]. Elevated or increasing serum MMP-3 levels in patients with RA are associated with the rapid progression of joint destruction. Conversely, serum MMP-3 levels decrease when the condition stabilizes in response to the therapeutic effect of DMARDs, including bDMARDs [13]. There are sex differences in serum MMP-3 levels; the upper limits of the reference range are 121.0 and 59.7 ng/mL for male and female individuals, respectively. Researchers have reported that normal serum MMP-3 levels, in combination with CRP levels or disease activity, are useful for predicting clinical remission and normal physical function in patients with RA [14]. However, progression of joint destruction is observed even in patients with RA with negative CRP values, disease activity in remission, or LDA [15]. Therefore, we observed patients with RA with negative CRP values during MTX monotherapy or combination therapy for 1 year and investigated whether serum MMP-3 levels are correlated with joint destruction. Furthermore, in this study, we developed an ideal cutoff value for MMP-3 based on the relationship between blood MMP-3 levels and joint destruction.

## METHODS

### Study design and patient selection

The retrospective study enrolled patients who visited Hyogo Medical University Hospital or Kyoto University Hospital between April 1, 2011 to April 30, 2021 and met the 1987 and/or 2010 RA classification criteria. Patients with RA (n = 182) whose CRP values became negative following MTX monotherapy or DMARD combination therapy (MTX together with other DMARDs) were divided into two groups based on their serum MMP-3 positivity at the end of the observation period, and the 1-year progression of joint destruction was retrospectively compared through X-rays.

### Assessment

The medical records of the patients, including information related to the visual analog scale (VAS), CRP, erythrocyte sedimentation rate (ESR), rheumatoid factor (RF), serum MMP-3, anti-citrullinated protein antibody (ACPA), 28 joint disease activity score (DAS28), and clinical disease activity index (CDAI), were retrospectively reviewed by two authors (HK, MM). Patients who had moderate or severe renal dysfunction (eGFRcre < 45 mL/min/1.73 m^2^) or were taking oral PSL > 25 mg/day were excluded. After the observation period, the patients were divided into the serum MMP-3 positive (MMP-3(+)) and MMP-3 negative (MMP-3(−)) groups based on whether their serum MMP-3 levels were above or below the upper reference limit for each sex. Progression was assessed via radiography of the hands, wrists, and feet and scored chronologically using the mTSS method recommended by Bruynesteyn et al. [16]. Radiographs of each patient’s hands and feet were taken upon the initiation of MTX therapy and 1 year after MTX therapy. Radiographic progression was evaluated independently by two rheumatologists (MM and KMurakami), who were trained and certified by Prof. van der Heijde (Leiden University) for the mTSS scoring system. Moreover, mTSS progression after 1 year (ΔmTSS) was calculated from the mean progression determined by the two readers. If ΔmTSS differed by ≥10, the two rheumatologists discussed and reached a consensus. Patients were classified as having structural remission (ΔmTSS < 0.5) or radiographic evidence of progression (ΔmTSS ≥ 0.5).

### Ethical approval

The retrospective study protocol was approved by the Ethics Committee of Hyogo Medical University (Protocol No. 3923). This retrospective study waived the requirement for individual informed consent owing to the “opt-out” principle. Patients were allowed to “opt-out” of the database if they wished. Clinical data at Kyoto University were approved by the Medical Ethics Committee of the Kyoto University Graduate School and Faculty of Medicine (No. R0357), and written informed consent to participate in the study was obtained from all patients of Kyoto University Hospital.

### Statistical analysis

All statistical analyses were performed using IBM SPSS Statistics version 29 (IBM Corp., Armonk, NY, USA). Fisher’s exact test was used for categorical variables. Receiver operating characteristic (ROC) curve analysis was used to determine the cutoff value and p-values of <0.05 were considered statistically significant.

## RESULTS

### Clinical characteristics of patients upon the initiation of MTX therapy

Of the patients who continued MTX monotherapy or DMARD combination therapy for at least 1 year, 182 (including 142 females) could be evaluated radiographically. Upon initiation of MTX therapy, participants had a mean age of 59.1 ± 12.1 years, with a mean disease duration of 9.3 ± 10.1 years. Among them, 64.2% were RF-positive, 64.2% were ACPA-positive, and 56.0% were both RF- and ACPA-positive. The mean DAS28-ESR was 2.46 ± 0.98, the mean CDAI was 4.51 ± 5.48, the mean MTX dose was 7.0 ± 2.7 mg/week in 125 (68.7%) patients, and the mean PSL dose was 4.1 ± 2.6 mg/day in 76 patients (41.8%) (Table 1).

**Table 1.**
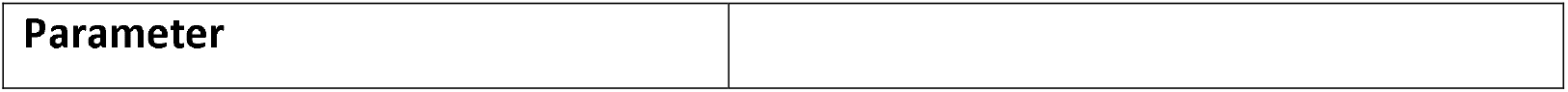
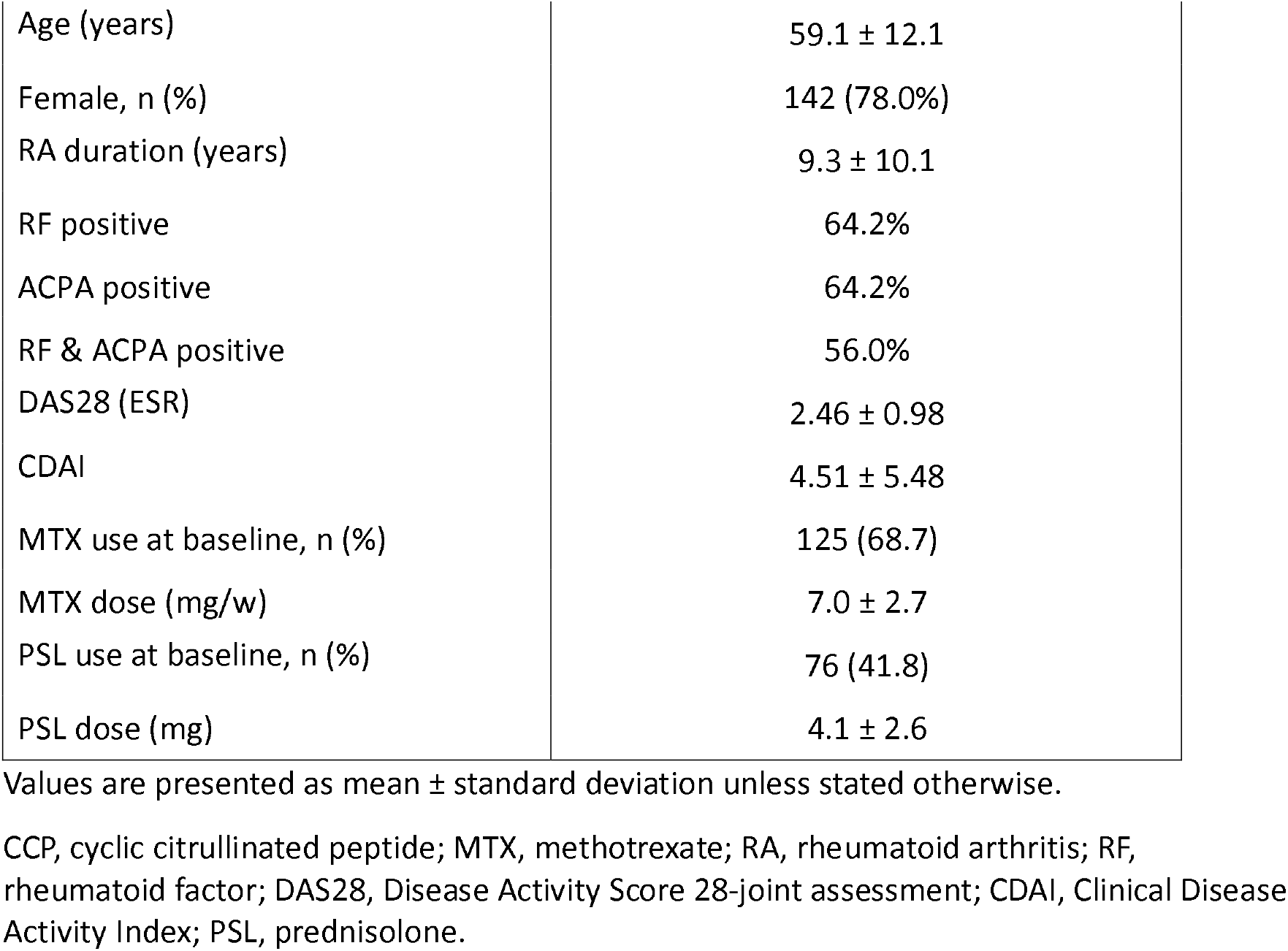
Subject demographics and clinical characteristics at 0week.

|  |  |
| --- | --- |
| Age (years) | 59.1 ± 12.1 |
| Female, n (%) | 142 (78.0%) |
| RA duration (years) | 9.3 ± 10.1 |
| RF positive | 64.2% |
| ACPA positive | 64.2% |
| RF & ACPA positive | 56.0% |
| DAS28 (ESR) | 2.46 ± 0.98 |
| CDAI | 4.51 ± 5.48 |
| MTX use at baseline, n (%) | 125 (68.7) |
| MTX dose (mg/w) | 7.0 ± 2.7 |
| PSL use at baseline, n (%) | 76 (41.8) |
| PSL dose (mg) | 4.1 ± 2.6 |

Values are presented as mean ± standard deviation unless stated otherwise.
CCP, cyclic citrullinated peptide; MTX, methotrexate; RA, rheumatoid arthritis; RF, rheumatoid factor; DAS28, Disease Activity Score 28-joint assessment; CDAI, Clinical Disease Activity Index; PSL, prednisolone.

### Comparison of the progression of joint damage estimated by radiography in the MMP-3(−) and MMP-3(+) groups

The progression rates of joint destruction in the MMP-3(−) (n = 100) and MMP-3(+) groups (n = 82) were 22.5% (n = 23) and 42.1% (n = 35), respectively (p < 0.01). Changes in the mTSS from baseline (ΔmTSS) were 0.46 ± 1.12 and 0.97 ± 1.99 (p = 0.03) (Fig. 1A), and the nonprogression rates (ΔmTSS < 0.5) were 77.5% and 57.9%, respectively (Fig. 1B). At baseline, the PSL usage rates were 26.0% (n = 26) and 61.0% (n = 50). The progression of joint destruction was examined in 106 of these 182 patients (MMP-3(−), n = 74; MMP-3(+), n = 32) who were PSL-free. Among these patients, the progression rates of joint destruction were 23.6% (n = 18) and 48.4% (n = 16) in the MMP-3(−) and MMP-3(+) groups, respectively (p = 0.01). The ΔmTSS values were 0.53 ± 1.24 and 1.16 ± 1.98 (p = 0.05) (Fig. 1C), and the nonprogression rates were 76.4% and 51.6%, respectively (Fig. 1D).

**Fig 1.** Progression of joint destruction was analyzed in the MMP-3(−) and MMP-3(+) groups at 52weeks. (A) Patients with rheumatoid arthritis (n = 183), change from baseline in the modified total sharp score (ΔmTSS). (B) Patients with rheumatoid arthritis (n = 183), cumulative probability plot of mTSS. (C) Patients with PSL-free rheumatoid arthritis (n = 106), change from baseline in ΔmTSS. (D) Patients with PSL-free rheumatoid arthritis (n = 106), cumulative probability plot of mTSS. (E) Patients with no swollen joints at 52 weeks (n = 97), change from baseline in the modified total sharp score (ΔmTSS). (F) Patients with no swollen joints at 52 weeks (n = 97), cumulative probability plot of mTSS. (G) Correlations between CRP and MMP-3 values in 182 patients. Values in (A)(C)(E) indicate the mean (SD) at each time point and in the MMP-3(−) or MMP-3(+) group. Percentages in (B)(D)(F) indicate the progression rates of joint destruction (ΔmTSS > 0.5) in the MMP-3(−) or MMP-3(+) group.

Even when the CRP value becomes negative at 52 weeks, it does not necessarily indicate the absence of inflammation. We sometimes encounter patients with a few small joint swellings and negative CRP values. Thus, we examined the progression of joint destruction in 97 of these 182 patients with no swollen joints at 52 weeks (MMP-3(−), n = 58; MMP-3(+), n = 39). Among these patients, the progression rates of joint destruction were 19.8% (n = 12) and 42.3% (n = 17) in the MMP-3(−) and MMP-3(+) groups, respectively (p = 0.02). The ΔmTSS values were 0.37 ± 0.89 and 1.18 ± 2.45 (p = 0.02) (Fig. 1E), and the nonprogression rates were 80.2% and 57.7%, respectively (Fig. 1F). The PSL usage rates at baseline were 27.6% (n = 16) and 53.8% (n = 21), respectively. We found no correlation between CRP and MMP-3 in 182 patients (Fig. 1G); therefore, joint destruction progresses more likely in the MMP-3(+) group than in the MMP-3(−) group (Table 2).

**Table 2.**
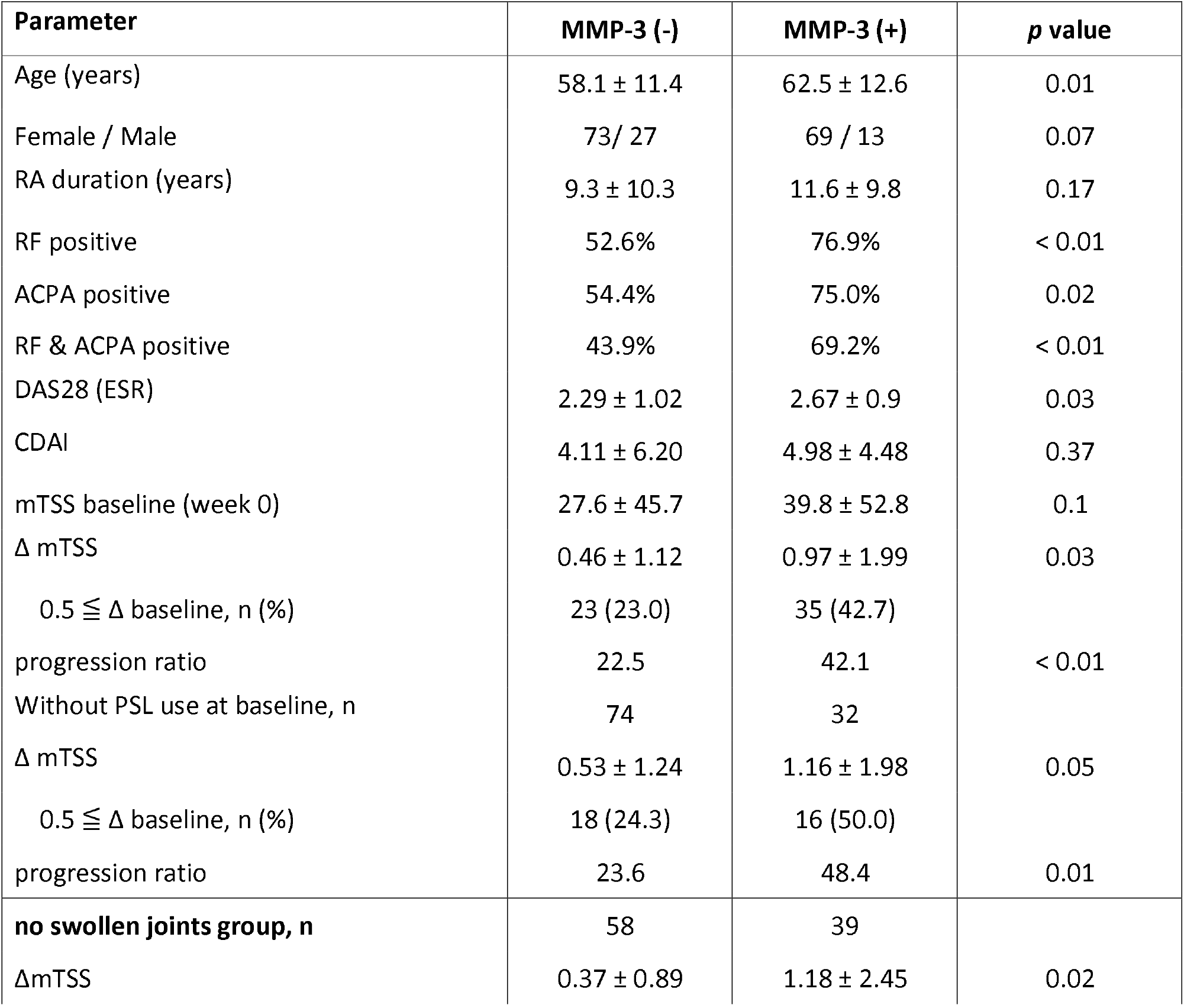
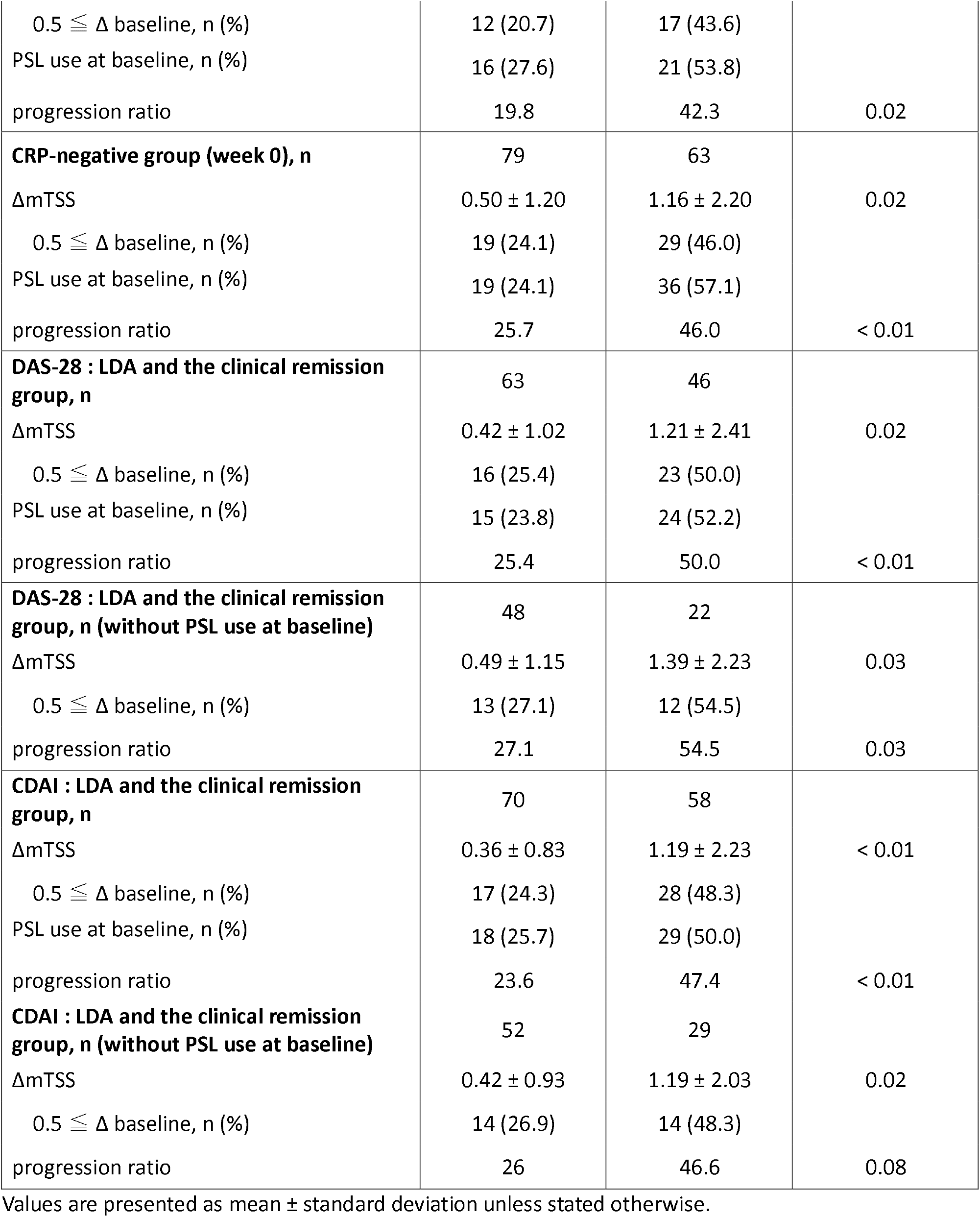
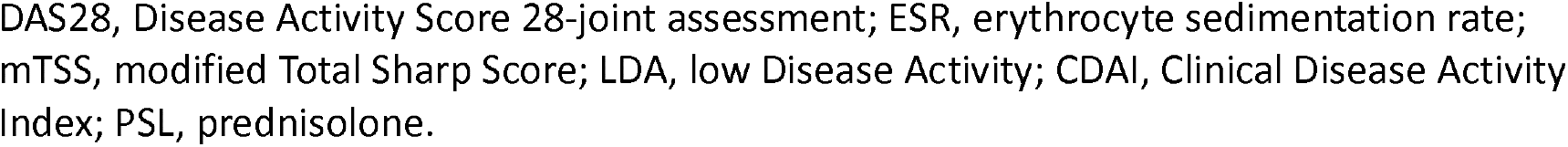
MMP-3 (-) group or MMP-3 (+) group analysis at 52weeks.

| Parameter | MMP-3 (-) | MMP-3 (+) | p value |
| --- | --- | --- | --- |
| Age (years) | 58.1 ± 11.4 | 62.5 ± 12.6 | 0.01 |
| Female / Male | 73/ 27 | 69 / 13 | 0.07 |
| RA duration (years) | 9.3 ± 10.3 | 11.6 ± 9.8 | 0.17 |
| RF positive | 52.6% | 76.9% | < 0.01 |
| ACPA positive | 54.4% | 75.0% | 0.02 |
| RF & ACPA positive | 43.9% | 69.2% | < 0.01 |
| DAS28 (ESR) | 2.29 ± 1.02 | 2.67 ± 0.9 | 0.03 |
| CDAI | 4.11 ± 6.20 | 4.98 ± 4.48 | 0.37 |
| mTSS baseline (week 0) | 27.6 ± 45.7 | 39.8 ± 52.8 | 0.1 |
| $\Delta$ mTSS | 0.46 ± 1.12 | 0.97 ± 1.99 | 0.03 |
| 0.5 ≤ $\Delta$ baseline, n (%) | 23 (23.0) | 35 (42.7) | |
| progression ratio | 22.5 | 42.1 | < 0.01 |
| Without PSL use at baseline, n | 74 | 32 |  |
| $\Delta$ mTSS | 0.53 ± 1.24 | 1.16 ± 1.98 | 0.05 |
| 0.5 ≤ $\Delta$ baseline, n (%) | 18 (24.3) | 16 (50.0) | |
| progression ratio | 23.6 | 48.4 | 0.01 |
| **no swollen joints group, n** | **58** | **39** |  |
| ΔmTSS | 0.37 ± 0.89 | 1.18 ± 2.45 | 0.02 |
| 0.5 ≤ $\Delta$ baseline, n (%) | 12 (20.7) | 17 (43.6) | |
| PSL use at baseline, n (%) | 16 (27.6) | 21 (53.8) |  |
| progression ratio | 19.8 | 42.3 | 0.02 |
| **CRP-negative group (week 0), n** | 79 | 63 |  |
| ΔmTSS | 0.50 ± 1.20 | 1.16 ± 2.20 | 0.02 |
| 0.5 ≤ $\Delta$ baseline, n (%) | 19 (24.1) | 29 (46.0) | |
| PSL use at baseline, n (%) | 19 (24.1) | 36 (57.1) |  |
| progression ratio | 25.7 | 46.0 | < 0.01 |
| **DAS-28 : LDA and the clinical remission group, n** | 63 | 46 |  |
| ΔmTSS | 0.42 ± 1.02 | 1.21 ± 2.41 | 0.02 |
| 0.5 ≤ $\Delta$ baseline, n (%) | 16 (25.4) | 23 (50.0) | |
| PSL use at baseline, n (%) | 15 (23.8) | 24 (52.2) |  |
| progression ratio | 25.4 | 50.0 | < 0.01 |
| **DAS-28 : LDA and the clinical remission group, n (without PSL use at baseline)** | 48 | 22 |  |
| ΔmTSS | 0.49 ± 1.15 | 1.39 ± 2.23 | 0.03 |
| 0.5 ≤ $\Delta$ baseline, n (%) | 13 (27.1) | 12 (54.5) | |
| progression ratio | 27.1 | 54.5 | 0.03 |
| **CDAI : LDA and the clinical remission group, n** | 70 | 58 |  |
| ΔmTSS | 0.36 ± 0.83 | 1.19 ± 2.23 | < 0.01 |
| 0.5 ≤ $\Delta$ baseline, n (%) | 17 (24.3) | 28 (48.3) | |
| PSL use at baseline, n (%) | 18 (25.7) | 29 (50.0) |  |
| progression ratio | 23.6 | 47.4 | < 0.01 |
| **CDAI : LDA and the clinical remission group, n (without PSL use at baseline)** | 52 | 29 |  |
| ΔmTSS | 0.42 ± 0.93 | 1.19 ± 2.03 | 0.02 |
| 0.5 ≤ $\Delta$ baseline, n (%) | 14 (26.9) | 14 (48.3) | |
| progression ratio | 26 | 46.6 | 0.08 |

Values are presented as mean ± standard deviation unless stated otherwise.
DAS28, Disease Activity Score 28-joint assessment; ESR, erythrocyte sedimentation rate; mTSS, modified Total Sharp Score; LDA, low Disease Activity; CDAI, Clinical Disease Activity Index; PSL, prednisolone.

### Effect of PSL-induced elevation of serum MMP-3 on the progression of joint destruction

In the MMP-3(+) group, progression of joint destruction was assessed in groups with (n = 46) or without (n = 36) PSL. We evaluated whether the oral PSL-induced serum MMP-3 increase was relevant to the joint destruction progression. However, progression of joint destruction was observed in 43.1% (n = 16) and 40.2% (n = 19) of the patients in the PSL (−) and PSL (+) groups (p = 0.82); ΔmTSS values were 1.03 ± 1.90 and 0.92 ± 2.08 (p = 0.82) (Fig. 2A); and the nonprogression rates were 56.9% and 59.7%, respectively (Fig. 2B). We found no significant difference in disease activity between the two groups (Table 3). These data suggest that the increase in serum MMP-3 levels induced by PSL is also involved in the progression of joint destruction.

**Table 3.** In the MMP-3 (+) group, PSL (-) group or PSL (+) group analysis at 52weeks.

| Parameter | PSL (-) | PSL (+) | *p* value |
| --- | --- | --- | --- |
| MMP-3 (+) group, n | 36 | 46 |  |
| DAS28 (ESR) | 2.49 ± 0.86 | 2.88 ± 1.02 | 0.11 |
| CDAI | 3.64 ± 3.23 | 5.73 ± 5.05 | 0.06 |
| ΔmTSS | 1.03 ± 1.90 | 0.92 ± 2.08 | 0.82 |
| 0.5 ≤ Δ baseline, n (%) | 16 (44.4) | 19 (43.6) |  |
| progression ratio | 43.1 | 40.2 | 0.82 |

Values are presented as mean ± standard deviation unless stated otherwise.
DAS28, Disease Activity Score 28-joint assessment; ESR, erythrocyte sedimentation rate; mTSS, modified Total Sharp Score; CDAI, Clinical Disease Activity Index; PSL, prednisolone.

**Fig 2.** For the MMP-3+ group (n = 83), progression of joint destruction was analyzed in the PSL(−) and PSL(+) groups at 52 weeks. (A) Change from baseline in the modified total sharp score (ΔmTSS). Values in indicate mean (SD) at each time point and in the PSL(−) or PSL(+) group. (B) Cumulative probability plot of mTSS. Percentages indicate the progression rates of joint destruction (ΔmTSS > 0.5) of each treatment group.

### Comparison of the progression of joint damage estimated by radiography between the MMP-3(−) and MMP-3(+) subgroups within the CRP-negative group at baseline

We examined the progression of joint destruction in 142 of 183 patients with RA (MMP-3(−), n = 79; MMP-3(+), n = 63) who were CRP negative at baseline. Progression of joint destruction was found in 25.7% and 46.0% (p < 0.01); the ΔmTSS values were 0.50 ± 1.20 and 1.16 ± 2.20 (p = 0.02) (Fig. 3A); and the nonprogression rates (ΔmTSS < 0.5) were 74.3% and 54.0% (Fig. 3B. The PSL usage rates at baseline were 24.1% (n = 19) and 57.1% (n = 36). This suggests that progression of joint destruction is likely if serum MMP-3 levels remain positive, even when baseline CRP is negative (Table 2).

**Fig 3.** For the CRP-negative group at baseline (n = 142), progression of joint destruction was analyzed in the MMP-3(−) and MMP-3(+) groups at 52 weeks. (A) Change from baseline in the modified total sharp score (ΔmTSS). Values indicate mean (SD) at each time point and MMP-3(−) or MMP-3(+) group. (B) Cumulative probability plot of mTSS. Percentages indicate the progression rates of joint destruction (ΔmTSS > 0.5) of each treatment group.

### Comparison of the progression of joint damage estimated by radiography in the MMP-3(−) and MMP-3(+) groups with LDA or clinical remission (DAS28-ESR < 2.6 or CDAI ≦ 2.8)

Clinical remission is the therapeutic target for patients with RA, with LDA being the best possible alternative. Thus, the study enrolled 109 of 183 patients with RA (MMP-3(−), n = 63; MMP-3(+), n = 46) who had achieved LDA or clinical remission (DAS28-ESR < 2.6). The progression of joint destruction was found in 24.6% (MMP-3(−)) and 48.9% (MMP-3(+)) (p < 0.01); the ΔmTSS values were 0.42 ± 1.02 and 1.21 ± 2.41 (p = 0.02) (Fig. 4A); and the nonprogression rates (ΔmTSS < 0.5) were 75.4% and 51.1%, respectively (Fig. 4B). The progression of joint destruction was further examined in 70 of these 109 patients (MMP-3(−), n = 48; MMP-3(+), n = 22) who were PSL-free. Among these patients, progression of joint destruction was observed in 26.0% and 47.7% (p = 0.03), the ΔmTSS values were 0.49 ± 1.15 and 1.39 ± 2.23 (p = 0.03), and the nonprogression rates were 72.9% and 45.5%, respectively. Similarly, 128 of 182 patients with RA (MMP-3(−), n = 70; MMP-3(+), n = 58) who had achieved LDA or clinical remission (CDAI ≦ 2.8) were compared. The progression of joint destruction was found in 23.6% (MMP-3(−)) and 47.4% (MMP-3(+)) (p < 0.01), the ΔmTSS values were 0.36 ± 0.83 and 1.19 ± 2.23 (p < 0.01) (Fig. 4C), and the nonprogression rates (ΔmTSS < 0.5) were 76.4% and 52.6%, respectively (Fig. 4D). We further examined the progression of joint destruction in 81 of these 128 patients (MMP-3(−), n = 52; MMP-3(+), n = 29) who were PSL-free. Among these patients, we observed progression of joint destruction in 26.0% and 46.6% (p = 0.08); the ΔmTSS values were 0.42 ± 0.93 and 1.19 ± 2.03 (p = 0.02); and the nonprogression rates were 74.0% and 53.4%, respectively. Joint destruction was more severe in the MMP-3(+) group than in the MMP-3(−) group (Table 2), indicating that neither the achievement of LDA nor clinical remission is sufficient to prevent the progression of joint destruction, particularly when the serum MMP-3 levels remain positive.

**Fig 4.** For patients with rheumatoid arthritis in remission or with low disease activity, progression of joint destruction was analyzed in the MMP-3(−) and MMP-3(+) groups at 52 weeks. (A) Patients with rheumatoid arthritis in remission or with low disease activity on the DAS28 (n = 109), change from baseline in the modified total sharp score (ΔmTSS). (B) Patients with rheumatoid arthritis in remission or with low disease activity on the DAS28 (n = 109), cumulative probability plot of mTSS. (C) Patients with rheumatoid arthritis in remission or with low disease activity on the CDAI (n = 128), change from baseline in ΔmTSS. (D) Patients with rheumatoid arthritis in remission or with low disease activity on the CDAI (n = 128), cumulative probability plot of mTSS. Values in (A)(C) indicate mean (SD) at each time point and in the MMP-3(−) or MMP-3(+) groups. Percentages in (B)(D) indicate the progression rates of joint destruction (ΔmTSS > 0.5) in the MMP-3(−) or MMP-3(+) group.

### ROC analysis to determine the cutoff value of serum MMP-3 levels

Finally, the optimal serum MMP-3 cutoff value was examined from the ROC curve using the Youden index. PSL-free female patients were selected because serum MMP-3 levels are affected by sex (the upper limits of the currently used reference range for male and female are 121.0 and 59.7 ng/mL, respectively) and PSL; the study included a few male patients. The ROC analysis revealed that the cutoff value of serum MMP-3 level was 49.7 ng/mL (area under the curve, 0.681; 95% CI 0.560–0.802, and p < 0.01) (Fig. 5A) to achieve structural remission (ΔmTSS < 0.5). In patients with advanced joint destruction (ΔmTSS ≥ 0.5), few patients had serum MMP-3 levels <49.7 ng/mL (Fig. 5B). Therefore, in female patients with RA, lowering the serum MMP-3 level to <49.7 ng/mL is desirable to prevent the progression of joint destruction.

**Fig 5.** ROC analysis to determine the cutoff value of serum MMP-3 levels. (A) The ROC analysis revealed the cutoff value of the serum MMP-3 was 49.7 ng/ml (area under the curve: 0.681, 95%CI: 0.560–0.802, p < 0.01) to achieve ΔmTSS < 0.5. (B) In the group of patients with advanced joint destruction (ΔmTSS ≥ 0.5), patients had serum MMP-3 levels more than 49.7 ng/ml.

## DISCUSSION

Progressive joint destruction is often observed during RA treatment, despite negative CRP values [7]. The authors of a previous study proposed that residual MMP-3 activity is one of the responsible factors [17]. Our study showed a greater progression of joint destruction in the MMP-3(+) group than in the MMP-3(−) group, despite arthritis being well controlled by MTX therapy. This was evidenced by the negative CRP values and the absence of swollen joints at 52 weeks. Furthermore, we found no correlation between CRP and MMP-3 levels. These results suggest that persistently high serum MMP-3 levels lead to joint destruction in patients with RA, including those in remission or with LDA.

During RA treatment, PSL administration is considered to provide a good therapeutic effect when used in appropriate doses [6]; however, serum MMP-3 levels tend to increase via an unknown mechanism [18, 19]. If this PSL-induced elevation of serum MMP-3 is simply affected by the detection of serum MMP-3 in the clinical laboratory and is not related to the pathophysiology of RA, joint destruction should progress more in the PSL(−) group than in the PSL(+) group when their MMP-3 values are the same. However, our results showed that the PSL-treated group had the same mean ΔmTSS and progression rates of joint destruction as the PSL-free group. To exclude the possibility that PSL was administered to patients with high disease activity, we investigated whether there was a correlation between disease activity and PSL usage. We found no significant differences in disease activity between the PSL (−) and PSL (+) groups. Furthermore, we observed no correlation between the PSL dose and the serum MMP-3 level. These data suggest that the PSL-induced increase in serum MMP-3 levels is also involved in the progression of joint destruction. To the best of our knowledge, this is the first report to show that a PSL-induced increase in serum MMP-3 levels is involved in joint destruction.

Interestingly, while we selected patients in remission or with LDA at baseline, the MMP-3(+) group had significantly higher mean ΔmTSS and progression rates of joint destruction than the MMP-3(−) group. Although it has been reported that joint destruction is correlated with disease activity [20], our results suggest that serum MMP-3 values are more important compared to DAS28-ESR and CDAI as an indicator of the radiographic progression of joint destruction. There was no correlation between DAS28 and MMP-3 or between CDAI and MMP-3 in patients with LDA or remission (data not shown). These results also suggest that serum MMP-3 levels serve as an indicator of joint destruction, which implies that even with reduced inflammation and improved disease activity to LDA or remission, joint destruction progression may still occur unless serum MMP-3 levels are appropriately managed.

Previous studies have reported cutoff values for serum MMP-3 levels to prevent joint destruction [21,22]. These studies included RA patients with positive CRP values, high disease activity, and receiving PSL treatment. Furthermore, these studies calculated the cutoff value from a patient cohort comprising both sexes, despite different upper limit values of the reference range. In the present study, we analyzed the cutoff values for PSL-free female patients with RA, negative CRP values, and remission or LDA. In the ROC analysis, the appropriate cutoff value of serum MMP-3 levels was found to be 49.7 ng/mL for preventing the progression of joint destruction in female patients; this value falls below the upper limit of the reference range (59.7 ng/mL). Serum MMP-3 levels may be a better indicator of whether joint destruction is likely to progress and suggest treatment intensification, even if disease activity and CRP levels have remained low for up to 1 year. The current reference range of serum MMP-3 was determined based on reference individuals who met the following conditions: negative CRP values, negative RF values, normal fasting blood sugar levels, and normal liver enzyme test results. However, these conditions do not exclude potential RA patients. Thus, it is not surprising that our cutoff value was lower than the upper limit of the reference range of serum MMP-3[23].

MMP-3 can activate several other MMPs, including MMP-1, MMP-7, and MMP-9, thereby increasing connective tissue matrix proteolysis [24]. It is thought that these proteases contribute to joint destruction, either directly or indirectly, by degrading the cartilage extracellular matrix [9, 11]. Thus, it is difficult to achieve structural remission unless serum MMP-3 levels are reduced to an appropriate level. However, there are currently no therapeutic agents that directly lower serum MMP-3 levels. In our in vitro experiment, TNF- alpha-induced increase in MMP-3 levels from RA synoviocytes was inhibited by PSL (data not shown); however, oral PSL administration increases serum MMP-3 levels in vivo [19]. In order to develop new treatments that can decrease the serum MMP-3 levels to prevent joint destruction in RA, it is necessary to elucidate the mechanism of the PSL-induced increase in serum MMP-3 levels or develop new therapeutic agents that lower MMP-3 levels.

### Limitations

This study has some limitations. The retrospective design limits the ability to draw causal inferences. The sample size is relatively small, and the findings may not be generalizable. The sample is predominantly from two university hospitals in the Kansai Area, Japan, which may limit generalizability. The number of male patients was too small to determine the adequate cutoff value of serum MMP-3 levels to prevent the progression of joint destruction in males. Joint destruction lasting longer than 52 weeks was not estimated.

## Conclusion

Our study indicates that residual MMP-3 activity, regardless of PSL administration, may lead to the progression of joint destruction in RA patients even after achieving clinical remission or LDA through successful treatment with MTX. Intensification of therapy may be necessary to achieve structural remission in female RA patients with serum MMP-3 levels of 49.7 ng/ml or higher.

## Data Availability

All relevant data are within the manuscript and its Supporting Information files.

## Acknowledgements

The authors thank the patients and medical staff of both hospitals for their contribution to this study. We would like to extend our heartfelt gratitude to Haruo Horii, Naohiro Ito, and Masatoshi Fujii for their generous financial support for the KURAMA cohort. The funders were not involved in the study design, data collection, analysis, interpretation of the data, writing of the manuscript, or in any decision to publish the results. We also thank Enago (www.enago.jp) for the English language review.

## REFERENCES

1. Koch AE (2007) The pathogenesis of rheumatoid arthritis. Am J Orthop (Belle Mead NJ) 36:5–8

2. Smolen JS, Aletaha D, Bijlsma JWJ et al (2010) Treating rheumatoid arthritis to target: Recommendations of an international task force. Ann Rheum Dis 69:631–637. 10.1136/ard.2009.123919

3. Smolen JS, Breedveld FC, Burmester GR et al (2016) Treating rheumatoid arthritis to target: 2014 update of the recommendations of an international task force. Ann Rheum Dis 75:3–15. 10.1136/annrheumdis-2015-207524

4. Cronstein BN (2005) Low-dose methotrexatellJ: A mainstay in the treatment of rheumatoid arthritis. Pharmacol Rev 57:15914465. 10.1124/pr.57.2.3

5. Pincus T, Yazici Y, Sokka T et al (2003) Methotrexate as the “anchor drug” for the treatment of early rheumatoid arthritis. Clin Exp Rheumatol 21:S179–S185

6. Smolen JS, Landewé RBM, Bijlsma JWJ et al (2020) EULAR recommendations for the management of rheumatoid arthritis with synthetic and biological disease-modifying antirheumatic drugs: 2019 update. Ann Rheum Dis 79:S685–S699. 10.1136/annrheumdis-2019-216655

7. Smolen JS, Van Der Heijde DMFM, St.Clair EW et al (2006) Predictors of joint damage in patients with early rheumatoid arthritis treated with high-dose methotrexate with or without concomitant infliximab: Results from the ASPIRE trial. Arthritis Rheum 54:702–710. 10.1002/art.21678

8. Malemud CJ (2006) Matrix metalloproteinases (MMPs) in health and disease: An overview. Front Biosci 11:1696–1701. 10.2741/1915

9. Yasuo Y, Hiroyuki N, Ken’ichi O et al (2000) Matrix metalloproteinases and tissue inhibitors ofmetalloproteinases in synovial fluids from patientswith rheumatoid arthritis or osteoarthritis. Ann Rheum Dis 59:455–461. 10.1136/ard.59.6.455

10. Cawston T (1998) Matrix metalloproteinases and TIMPs: Properties and implications for the rheumatic diseases. Mol Med Today 4:130–137. 10.1016/S1357-4310(97)01192-1

11. Okada Y (2000) Matrix-degrading metalloproteinases and their roles in joint destruction. Mod Rheumatol 10:121–128. 10.3109/s101650070018

12. Lerner A, Neidhöfer S, Reuter S, Matthias T (2018) MMP3 is a reliable marker for disease activity, radiological monitoring, disease outcome predictability, and therapeutic response in rheumatoid arthritis. Best Pract Res Clin Rheumatol 32:550–562. 10.1016/j.berh.2019.01.006

13. Kobayashi A, Naito S, Enomoto H et al (2007) Serum levels of matrix metalloproteinase 3 (stromelysin 1) for monitoring synovitis in rheumatoid arthritis. Arch Pathol Lab Med 131:563–570. 10.1043/1543-2165(2007)131

14. Hattori Y, Kida D, Kaneko A (2019) Normal serum matrix metalloproteinase-3 levels can be used to predict clinical remission and normal physical function in patients with rheumatoid arthritis. Clin Rheumatol 38:181–187. 10.1007/s10067-017-3829-9

15. Yoshimi R, Hama M, Takase K et al (2013) Ultrasonography is a potent tool for the prediction of progressive joint destruction during clinical remission of rheumatoid arthritis. Mod Rheumatol 23:456–465. 10.1007/s10165-012-0690-1

16. Bruynesteyn K, Boers M, Kostense P et al (2005) Deciding on progression of joint damage in paired films of individual patients: Smallest detectable difference or change. Ann Rheum Dis 64:179–182. 10.1136/ard.2003.018457

17. Konishi H, Kanou SE, Yukimatsu R et al (2022) Adenosine inhibits TNFα-induced MMP-3 production in MH7A rheumatoid arthritis synoviocytes via A2A receptor signaling. Sci Rep 12:1–9. 10.1038/s41598-022-10012-6

18. Hattori Y, Kida D, Kaneko A (2018) Steroid therapy and renal dysfunction are independently associated with serum levels of matrix metalloproteinase-3 in patients with rheumatoid arthritis. Mod Rheumatol 28:242–248. 10.1080/14397595.2017.1354431

19. Ribbens C, Martin y Porras M, Franchimont N, Kaiser M, Jaspar J, Damas P, Houssiau FMM (2002) Increased matrix metalloproteinase-3 serum levels in rheumatic diseases: Relationship with synovitis and steroid treatment. Ann Rheum Dis 61:161–166. 10.1136/ard.61.2.161

20. Tsuji H, Yano K, Furu M et al (2017) Time-averaged disease activity fits better joint destruction in rheumatoid arthritis. Sci Rep 7:1–8. 10.1038/s41598-017-05581-w

21. Mamehara A, Sugimoto T, Sugiyama D et al (2010) Serum matrix metalloproteinase-3 as predictor of joint destruction in rheumatoid arthritis, treated with non-biological disease modifying anti-rheumatic drugs. Kobe J Med Sci 56:98–107

22. Maksymowych WP, Landewé R, Conner-Spady B et al (2007) Serum matrix metalloproteinase 3 is an independent predictor of structural damage progression in patients with ankylosing spondylitis. Arthritis Rheum 56:1846–1853. 10.1002/art.22589

23. Yumeto N, Yuko T, Satoshi N et al (2014) Basic study of reagents for measurement of matrix metalloproteinase-3. Igakukensa 63:579–585. 10.1007/s12291-010-0025-y

24. Ogata Y, Enghild JJ, Nagase H (1992) Matrix metalloproteinase 3 (stromelysin) activates the precursor for the human matrix metalloproteinase 9. J Biol Chem 267:3581–3584. 10.1016/S0021-9258(19)50563-4

